# DBT-2026, a de-identified publicly available dataset of digital breast tomosynthesis exams with ground truth biopsies

**DOI:** 10.64898/2026.03.03.25337924

**Authors:** Jie Wu, Luiz Perandini, Tushar Batra, Sergei Igoshin, Sina Bari, Aline L. de Araujo, Martin J. Willemink

## Abstract

Digital breast tomosynthesis (DBT) is a powerful imaging modality that allows for improved lesion visibility, characterization, and localization compared to conventional two-dimensional digital mammography. DBT has been increasingly adopted in screening and diagnostic settings globally, particularly for women with dense breast tissue where tissue overlap presents a significant diagnostic challenge. Here we describe DBT-2026, a real world imaging dataset with 558 DBT exams from 558 patients with breast imaging reporting and data system (BI-RADS) scores of 0, 1, or 2. Each case contains one DBT examination in combination with expert annotations and free-text radiology reports that describe the radiological findings, produced in routine clinical practice. To protect patient privacy, all images and reports have been de-identified. The dataset is made freely available to researchers for non-commercial projects to facilitate and encourage research in breast cancer imaging.

## Introduction

With more than 39 million examinations per year, breast cancer screening is one of the most common tasks performed in radiological practice [1]. For this reason, it has been among the most studied medical imaging applications of artificial intelligence (AI). Mammography is the most common imaging modality studied within breast cancer screening. However, other modalities such as digital breast tomosynthesis (DBT), also referred to as three-dimensional (3D) mammography, are becoming more important. DBT is an advanced imaging modality designed to overcome some limitations of conventional two-dimensional (2D) digital mammography [2]. DBT acquires multiple low-dose projection images of the compressed breast at different angles over a limited arc, which are then reconstructed into thin, quasi-3D slices.

This technique reduces the impact of overlapping breast tissue, thereby improving lesion visibility, characterization, and localization. Clinical applications of DBT extend across both breast cancer screening and diagnostic imaging. Multiple clinical studies and large population-based trials have shown that DBT increases invasive cancer detection rates while simultaneously reducing false-positive recalls compared to 2D mammography alone [3–5]. For these reasons, DBT has been increasingly adopted in screening and diagnostic settings worldwide, particularly for women with dense breast tissue where tissue overlap presents a significant diagnostic challenge [6,7].

There is a lack of well-annotated publicly available DBT datasets with high quality ground truth outcome (biopsy). Most public datasets with DBT exams have relatively low numbers of patients with malignancies [1]. Here we describe DBT-2026, a real world imaging dataset of DBT screening exams from patients with breast imaging reporting and data system (BI-RADS) scores of 0, 1, or 2. The dataset is made freely available to researchers for non-commercial projects to facilitate and encourage research in breast cancer imaging.

## Materials & Methods

### Data curation

Requirement for individual patient consent was waived due to the retrospective nature of this project and all protected health information (PHI) was removed. DBT exams were sourced from an automated connection with the hospital picture archiving and communication system (PACS) in digital imaging and communications in medicine (DICOM) format through Segmed Piper (Segmed, Inc; Palo Alto, California). DICOM files contain images as well as metadata. Burnt-in text in images was evaluated using optical character recognition (OCR) technology that converts text on images into machine-readable text. Subsequently, text-based information (from both reports and images) was de-identified by removing the 18 Health Insurance Portability and Accountability Act (HIPAA) defined PHI identifiers from all text using a combination of natural language processing (NLP) and large language models (LLMs). Final images and text were manually evaluated by a clinical specialist (L.P.) and no PHI was detected.

A search using Segmed Openda (Segmed, Inc; Palo Alto, California) was performed for DBT exams obtained within the Midwest region of the United States of America. This cohort building tool allows for searching through modality, body part, radiology report, and DICOM metadata. Inclusion criteria included women who are 18 years or older who are eligible for breast cancer screening; bilateral exams that contain at least one mediolateral oblique (MLO) image and one craniocaudal (CC) for both breasts; no breast implants, previous breast/axilla surgery, or history of breast cancer. The search was narrowed to screening patients with BI-RADS scores of 0, 1, or 2. The dataset was enriched by prioritizing patients with biopsy-proven malignancies. Demographics (age, gender, race, ethnicity), BI-RADS score, biopsy results, and radiology report were exported to a comma separated values (CSV) document.

### Annotations

Lesion localization, characterization, segmentation, and measurement was performed by a team of fellowship-trained senior Breast Imaging Radiologists based in India, recruited and credentialed using the <u>iMerit Scholars</u> platform (iMerit Inc; San Jose, California). Radiologists followed a standardized annotation protocol developed by US-based clinical experts to ensure consistency in lesion categorization and measurement.

Data structuring and annotation were conducted using iMerit’s cloud-based, HIPAA-compliant <u>Ango Radiology Annotation Suite</u> (iMerit Inc; San Jose, California), which supports structured reporting, calibrated measurement tools, and audit-trailed version control. A two-step “Doer–Checker” workflow was implemented, whereby each case was independently reviewed for completeness and accuracy prior to finalization. Discrepancies were resolved through secondary review and consensus.

A final quality assurance review was performed by a U.S.-based, mammography quality standard act (MQSA)-certified breast imaging radiologist to validate lesion characterization and segmentation fidelity.

### Dataset publishing

The dataset is made available online (https://www.segmed.ai/resources/ai-for-impact-on-breast-cancer-dataset-form). Access is restricted to researchers who complete the registration and credentialing process and agree to the Data Use Agreement. The agreement specifies that the dataset may be used only for non-commercial research purposes, prohibits redistribution or sharing of the data with others, forbids any attempts at patient re-identification, and excludes any use for clinical decision-making. Each researcher must register individually and agree to these terms prior to accessing the dataset. All rights, title, and interest in the dataset remain with the authors and Segmed.

## Resulting dataset

Patient information is summarized in **Table 1**. A total of 558 DBT exams from 558 unique patients were included. The mean age was 57.5±11.1 years. The cohort was divided into four groups as summarized in **Table 2**. A total of 271 patients were biopsy-proven malignant (group A), 140 patients were benign (group B), 115 patients had a non-cancer callback (group C), and 32 patients were biopsy-proven benign (group D).

**Table 1:**
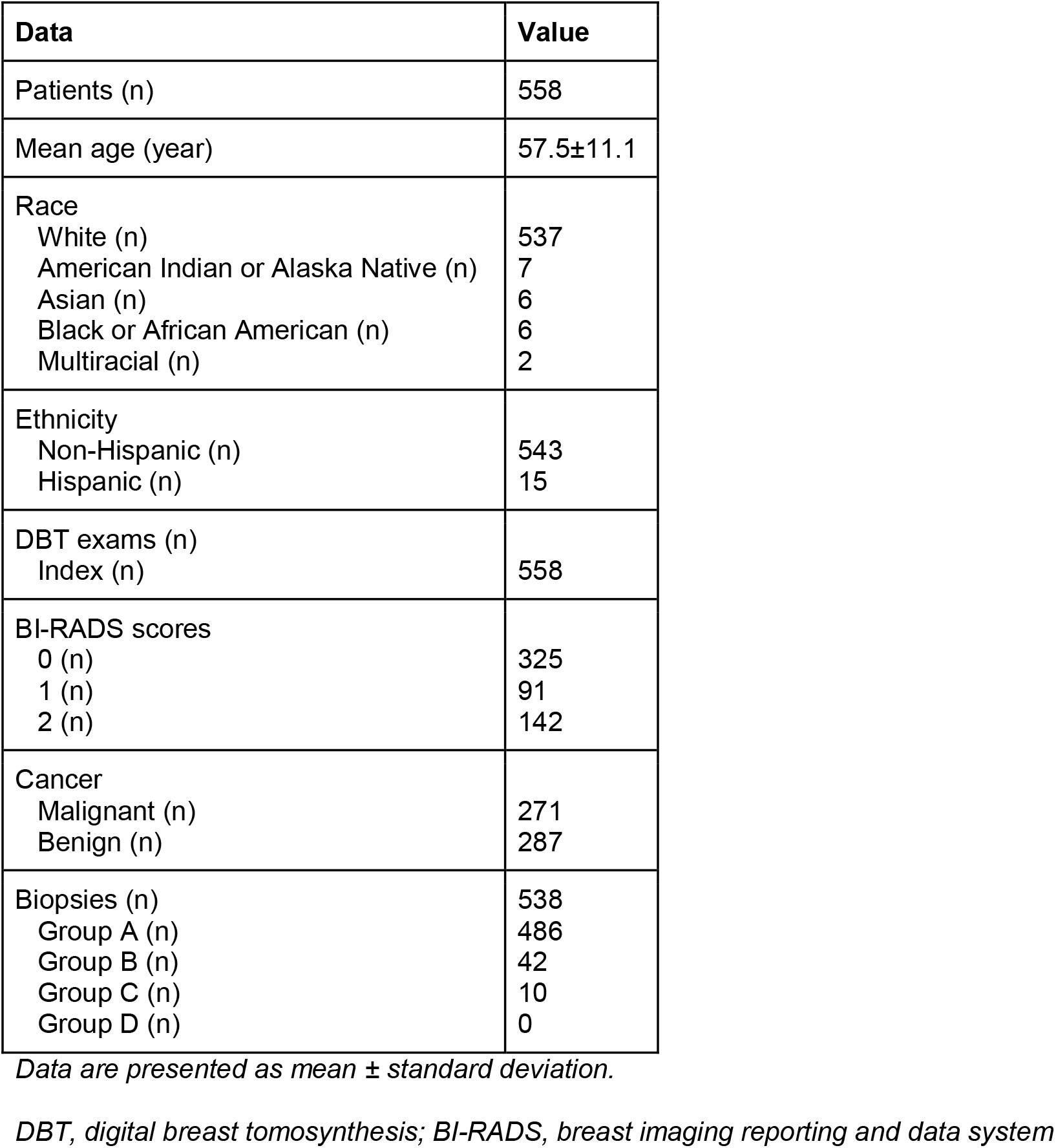
Descriptive statistics for DBT-2026 dataset.

**Table 2:** Distribution of the cohort.

| Group | Malignant/Benign | Description | Patients (n) |
| --- | --- | --- | --- |
| A | Malignant | Biopsy-proven malignant | 271 |
| B | Benign | Benign | 140 |
| C | Benign | Non-cancer callback screeners | 115 |
| D | Benign | Biopsy-proven benign | 32 |

## Discussion

We describe the curation of a dataset of 1,110 DBT exams from 558 patients. This dataset is unique since it contains high quality DBT exams with strong ground truth information (biopsy or follow-up). The dataset specifically addresses patients who received a BI-RADS score of 0 at screening, indicating an abnormality was found and additional examination was needed; and patients with relatively low BI-RADS scores of 1 or 2 based on imaging. Almost half of patients had a biopsy-proven malignancy (n=271, 48.6%).

A limitation is that all data is sourced from a single hospital and data cannot be used for commercial purposes. However, researchers can reach out to the authors to gain access to a more extensive multi-center database.

In summary, this publicly available dataset will contribute to the training and validation of AI models for breast cancer detection and characterization on DBT images. Permission for access to the database will be reviewed on a case-by-case basis.

## Data Availability

The dataset is made available online (https://www.segmed.ai/resources/ai-for-impact-on-breast-cancer-dataset-form).

## Acknowledgement

The authors would like to acknowledge Amy Sobel, MD, breast radiologist at Advocate Health for her contributions to the manuscript.

## Notes

### Competing Interest Statement

JW, LP, AL, and MJW are employees of Segmed. SB is employee at iMerit. No other relationships or activities that could appear to have influenced the submitted work.

### Funding Statement

This study did not receive any funding.

## References

1. Buda M, Saha A, Walsh R, Ghate S, Li N, Swiecicki A, et al. A Data Set and Deep Learning Algorithm for the Detection of Masses and Architectural Distortions in Digital Breast Tomosynthesis Images. JAMA Netw Open. 2021;4: e2119100.

2. Niklason LT, Christian BT, Niklason LE, Kopans DB, Castleberry DE, Opsahl-Ong BH, et al. Digital tomosynthesis in breast imaging. Radiology. 1997;205: 399–406.

3. Ciatto S, Houssami N, Bernardi D, Caumo F, Pellegrini M, Brunelli S, et al. Integration of 3D digital mammography with tomosynthesis for population breast-cancer screening (STORM): a prospective comparison study. Lancet Oncol. 2013;14: 583–589.

4. Skaane P, Bandos AI, Gullien R, Eben EB, Ekseth U, Haakenaasen U, et al. Comparison of digital mammography alone and digital mammography plus tomosynthesis in a population-based screening program. Radiology. 2013;267: 47–56.

5. Friedewald SM, Rafferty EA, Rose SL, Durand MA, Plecha DM, Greenberg JS, et al. Breast cancer screening using tomosynthesis in combination with digital mammography. JAMA. 2014;311: 2499–2507.

6. Rafferty EA, Durand MA, Conant EF, Copit DS, Friedewald SM, Plecha DM, et al. Breast Cancer Screening Using Tomosynthesis and Digital Mammography in Dense and Nondense Breasts. JAMA. 2016;315: 1784–1786.

7. McDonald ES, Oustimov A, Weinstein SP, Synnestvedt MB, Schnall M, Conant EF. Effectiveness of Digital Breast Tomosynthesis Compared With Digital Mammography: Outcomes Analysis From 3 Years of Breast Cancer Screening. JAMA Oncol. 2016;2: 737– 743.

